# Traditional Hemorrhoid Treatment Complications and Community Perspectives: Evidence from Southern Ethiopia

**DOI:** 10.64898/2026.07.09.26357622

**Authors:** Yidnekachew Mekonnen Bekele, Hunachew Beyene Mengesha, Tigabu Daniel Ayase, Abebe Melis Nisro

## Abstract

**Background:** Hemorrhoids are among the most common anorectal disorders, yet traditional treatment practices remain widespread in Ethiopia. These remedies often involve corrosive chemicals, herbal preparations, or invasive procedures, and are associated with severe complications. Despite their prevalence, systematic evidence on outcomes and community perceptions is limited.

**Methods:** A hospital-based cross-sectional study was conducted from December 30, 2024 to December 29, 2025 in Sidama, Ethiopia. A total of 450 patients diagnosed with hemorrhoids and managed across five government hospitals were enrolled. Structured questionnaires and medical record review were used to collect socio-demographic characteristics, clinical presentation, hospital management, traditional treatment practices, complications, and community perceptions. Descriptive statistics and independent sample t-tests were applied.

**Results:** The mean age of participants was 35.2 years, with a predominance of males (63.1%) and urban residents (72%). Perianal pain (84%) and rectal bleeding (50%) were the most frequent symptoms. Independent samples t-test analysis demonstrated that patients who visited traditional healers were significantly older than those who did not (mean age 48.2 vs. 34.4 years; mean difference = 13.8 years, 95% CI: 8.8–18.8; *p* < 0.001). Hospital management, primarily hemorrhoidectomy (31.8%), achieved favorable outcomes, with 97.3% of patients improving. Twenty-eight patients (6.2%) reported using traditional healers, most commonly involving topical chemical applications (71.4%). Complications were frequent among traditional users, with 85.7% experiencing adverse outcomes such as persistent pain, anal stenosis, and perianal discharge. Despite these complications, community perceptions remained largely positive or neutral, influenced by family and peers.

**Conclusion:** Traditional hemorrhoid treatment in Southern Ethiopia is associated with high complication rates, yet community perceptions remain favorable due to sociocultural influences. Hospital management demonstrates superior outcomes. Bridging the gap between biomedical care and community beliefs is essential to reduce morbidity and promote safe treatment.

## Introduction

Hemorrhoids are among the most frequent outpatient complaints. Patients usually present with rectal bleeding, itching and a protruding anal mass (1). Serious complications following traditional treatment have occurred but remain inadequately reported(2,3). Documented complications include anal stenosis, sepsis, bleeding, tetanus, hepatitis infection, and death. Anal stenosis is the narrowing of the anal opening that makes passage of stool difficult or impossible due to post-traditional medication applications or anorectal surgery(3–5).

In developing countries, most patients go to traditional healers for hemorrhoid treatment because they believe surgery will not cure hemorrhoids completely or fear recurrence(6,7). One study showed that hemorrhoids are the fourth most common disease treated by traditional medication in Ethiopia, although the real outcome is not yet determined(8).

Traditional healers often use corrosive substances such as acids, bases, and other toxins. These substances can cause mucosal irritation and necrosis. Anal stenosis, sepsis, incontinence, fistula, and tetanus are the common associated complications(9). Alongside these practices, medicinal plants are widely used to treat hemorrhoids, though adverse outcomes are rarely documented(10,11). Diverse plant families are applied, with shrubs and herbs being the most frequent forms(10,12,13).

In Africa, Ethiopia stands out for its extensive use of traditional medicine, hosting about 800 medicinal plant species commonly prepared from leaves, stems, roots, and barks(14,15). More than 90% of the population uses traditional remedies. Despite this reliance, perianal complications remain under-reported(8). Unlike India, where pilex extracts are standardized, Ethiopia lacks defined formulations(8,16). Reports from Kenya describe systemic side effects but not perianal complications(12).

Although some herbal medicines show therapeutic potential, most have not undergone clinical trials to assess serious adverse outcomes such as anal stenosis(12,15). Limited studies have been systematically explained these complications, despite physicians frequently observing them in practice(13,14,16). Furthermore, in Ethiopia and Nigeria, both patients and healers promote traditional medicine as an income source, contributing to under-reporting of negative effects(7).

These complications require extensive investigation to determine true rates and identify responsible substances. This study documented the prevalence and patterns of complications associated with traditional hemorrhoid treatment and can provide evidence to inform community awareness, clinical practice, and health policy.

## Methods

### Study area

Hawassa is a city in Ethiopia, on the shores of Lake Hawassa in the Great Rift Valley. It is 273 km south of Addis Ababa via Bishoftu, 130 km east of Sodo, and 75 km north of Dilla. The town serves as the capital of the Sidama Region. It lies on the Trans-African Highway 4, Cairo-Cape Town and has a latitude and longitude of 7°3′N 38°28′E and an elevation of 1,708 meters (5,604 ft) above sea level. Its name comes from a Sidamic word meaning “wide body of water”. Hawassa city has more than 13 hospitals. Hawassa city is one of largely populated city holding major traditional medication practicing traditional healers(17).

The other study area was Yirgalem town in southern Ethiopia, surrounded by Lake Woyima and Gidawo. It’s located 310 km south of Addis Ababa and 40 kilometers south of Hawassa in the Sidama region. This is another common site where traditional treatment is claimed to be used.

### Study Design and Period

A hospital-based cross-sectional study was conducted on patients with perianal complaints from December 30 2024 to December 29 2025.

#### Population, Eligibility, Sample size determination and sampling technique and procedure

The study population included all patients diagnosed with hemorrhoids and managed by treating physicians at Yirgalem General Hospital, Adare General Hospital, Tulla General Hospital, Motite Fura Primary Hospital, and Hawassa University comprehensive specialized hospitals. Patients were eligible if they had a confirmed diagnosis of hemorrhoids and received hospital management, regardless of whether they had previously sought traditional treatment. Patients who were unable to communicate, mentally ill, and severely ill were excluded from the study.

Since no prior study was available, a 50% complication rate among traditional medicine users was assumed. Using a single-proportion formula with 95% confidence level and 5% margin of error, the minimum sample size was calculated as 384. After adding 10% for non-response (38 participants), the final sample size was 422. A systematic sampling method was employed, and eligible patients presenting to the outpatient departments during the study period were approached and enrolled by trained data collectors.

### Study variables, operational definitions

The dependent variable in this study was the occurrence of complications following traditional hemorrhoid treatment. Independent variables included socio-demographic characteristics such as age, sex, marital status, occupation, residency, as well as clinical factors like type of hemorrhoid, presenting symptoms, prior diagnosis, and treatment history. Community perception variables were also considered, including beliefs about surgical cure, fear of recurrence, and attitudes toward traditional healers.

- **Traditional treatment user:** Patient reporting use of herbal remedies, corrosive chemicals, or other non-medical interventions for hemorrhoids.
- **Complication:** any adverse outcome following traditional treatment, including anal stenosis, sepsis, bleeding, tetanus, hepatitis infection, or death.
- **Community perception:** Community perception was defined as patient or caregiver views on effectiveness, safety, and recurrence risk of traditional hemorrhoid treatment, measured by asking respondents how the community perceives traditional healers.

### Data collection tools and procedures and data quality management

Data were collected using a structured questionnaire developed in Kobo Toolbox, designed to capture sociodemographic characteristics, clinical presentation, hospital treatment, traditional healer practices, and complication patterns. Trained data collectors interviewed patients attending outpatient departments to gather their perceptions, while additional information was abstracted from medical records. All responses were recorded directly into Kobo Toolbox. To ensure data quality, regular supervision and monitoring were conducted throughout the collection process. The research team reviewed entries daily for completeness and consistency, and any inconsistent or missing values were verified against patient records before analysis. Data cleaning was performed before export, and only validated datasets were used for statistical analysis.

### Data processing and analysis

Data collected through Kobo Toolbox were exported into SPSS version 26 for analysis. Before analysis, the dataset was cleaned and checked for completeness, consistency, and missing values verified against patient records. Descriptive statistics such as frequencies, percentages, means, and standard deviations were used to summarize sociodemographic characteristics, clinical features, and complication patterns. Independent sample t-test was used to compare mean age between patients who visited traditional healers and those who did not. Results were presented using text and tables to provide a clear overview of the findings.

### Ethical consideration

Ethical clearance and approval of the study were obtained from the Institutional Review Board (IRB) of Hawassa University College of Medicine and Health Sciences with reference number IRB/412/16. Written informed consent was obtained from all participants before each interview, with the consent form translated into the local language and read aloud when necessary. Participants were informed about the objectives and purpose of the study and were assured that they could withdraw at any time without consequence. Confidentiality was strictly maintained, and personal privacy and dignity were respected throughout the study. Clinical information was abstracted from patient medical records during the study period, with access limited to the research team, and no identifiable personal information was retained in the dataset used for analysis.

## Result

### Socio-demographic characteristics

A total of 450 patients diagnosed with hemorrhoids were included in the study. The mean age of participants was 35.2 ± 13.5 years, ranging from 16 to 84 years. Patients who visited traditional healers were significantly older than those who did not, with a mean age of 48.2 years compared to 34.4 years (mean difference = 13.8 years, 95% CI: 8.8–18.8, *p* < 0.001). The majority of patients were male (63.1%), and more than two-thirds (68%) were married. Most participants resided in urban areas (72%). In terms of occupation, government employees accounted for 33.8% of the study population (Table 1).

**Table 1.** Socio-demographic characteristics of study participants.

| Variables | Categories | Frequency | Percent (%) |
| --- | --- | --- | --- |
| **Sex** | Male | 284 | 63.1 |
|  | Female | 166 | 36.9 |
| **Residence** | Rural | 126 | 28 |
|  | Urban | 324 | 72 |
| **Marital status** | Married | 306 | 68 |
|  | Single | 139 | 30.9 |
|  | Widowed | 5 | 1.1 |
| **Occupation** | Farmer | 74 | 16.4 |
|  | Government employee | 152 | 33.8 |
|  | Merchant | 130 | 28.9 |
|  | Student | 76 | 16.9 |
|  | Others | 18 | 4 |

### Clinical presentation, hospital management, and treatment outcomes

The most common symptom was perianal pain, reported in 84% of patients. Only 9.1% of patients reported that they had previously been told they had hemorrhoids before their current hospital management. Regarding hospital management, 143 patients (31.8%) underwent hemorrhoidectomy. More advanced interventions were less frequent: 15 patients (3.3%) underwent dilation for anal stenosis.

Overall, hospital outcomes were favorable. The vast majority of patients (97.3%) improved following treatment (Table 2).

**Table 2.** Clinical presentation, hospital management, and treatment outcomes of study participants.

| Variable description |  | Frequency | Percent (%) |
| --- | --- | --- | --- |
| Patient presentation | Perianal pain | 378 | 84 |
|  | Bleeding per rectum | 225 | 50 |
|  | Failure to pass feces and flatus | 74 | 16.4 |
|  | Mass protruding per anus | 215 | 47.8 |
|  | Pruritus around anus | 151 | 33.6 |
|  | Perianal discharge | 50 | 11.1 |
|  | Skin tags | 22 | 4.9 |
| Prior diagnosis of hemorrhoid | Yes | 41 | 9.1 |
|  | No | 409 | 90.9 |
| Hospital management | Hemorrhoidectomy | 143 | 31.8 |
|  | Excision of thrombosed hemorrhoid | 84 | 18.7 |
|  | Dilation for anal stenosis | 15 | 3.3 |
|  | Diversion colostomy | 9 | 2 |
|  | Flap for anal stenosis | 5 | 1.1 |
|  | Conservative | 194 | 43 |
| Hospital treatment outcomes | Improved | 438 | 97.3 |
|  | Same | 11 | 2.4 |
|  | Worsening | 1 | .2 |

The most commonly diagnosed hemorrhoid was External hemorrhoid (57.1%), followed by internal hemorrhoid (27.1%) and interoexternal (15.8%). Among these 100% diagnosis was made by history and physical examination, while 3.8 % supplemented with colonoscopy

#### Traditional treatment users

A total of 28 patients (6.2%) reported receiving treatment from traditional healers. Sixteen had been previously diagnosed with hemorrhoids in hospitals and were on conservative management before seeking traditional remedies. Reported treatment modalities included topical application of unknown chemicals, ingestible preparations, and pricking with sharp materials.

Complications were frequent: 24 patients (85.7%) developed at least one adverse outcome, most commonly persistent perianal pain, perianal discharge, and anal stenosis. Hospital management of these complications involved hemorrhoidectomy, diversion colostomy, and flap procedures for anal stenosis (Table 3).

**Table 3.** Traditional treatment options provided and complications.

| Variable |  | Frequency (Yes) | Percent (%) |
| --- | --- | --- | --- |
| Treatment modalities | Pricking with sharp materials | 4 | 14.3 |
|  | Applying chemicals topically | 20 | 71.4 |
|  | Ingestible preparations | 11 | 39.3 |
| Complications | Any complication | 24 | 85.7 |
|  | Persistent perianal pain | 18 | 64.3 |
|  | Bleeding | 8 | 28.6 |
|  | Incontinence | 4 | 14.3 |
|  | Anal stenosis | 9 | 32.1 |
|  | Perianal discharge | 14 | 50 |

Hospital management of these complications involved hemorrhoidectomy (29.6%), diversion colostomy, and flap procedures for anal stenosis.

#### Community perception and sources of influence

Despite these adverse outcomes, community attitudes toward traditional treatment remained largely positive or neutral. Among the 28 patients, 12 (42.9%) reported a good perception, 10 (35.7%) reported a neutral perception, and 6 (21.4%) reported a negative perception.

Patients’ decisions to seek traditional healers were often influenced by others: 19 (67.9%) were advised by family members, 20 (71.4%) by previously treated individuals, while only 5 (17.9%) reported going on their own initiative.

## DISCUSSION

The majority of participants in this study were young to middle-aged adults (mean age 35.2 years). These findings are consistent with reports from Ethiopia, where hemorrhoids were more prevalent among working-age populations in Gondar(18). Similarly, a study from India found that hemorrhoids were more common in adults over 40 years, reflecting a comparable demographic pattern in South Asia(19).

Independent samples t-test analysis showed that patients who visited traditional healers were significantly older than those who did not (mean age 48.2 vs. 34.4 years, mean difference = 13.8 years, 95% CI: 8.8–18.8, *p* < 0.001). This finding underscores that reliance on traditional treatment was more entrenched among older individuals, who may be more influenced by cultural continuity and community traditions. Comparable evidence from Somalia demonstrated that older age groups (≥60 years) were significantly more likely to seek traditional healers, and those treated traditionally experienced higher complication rates(9). Despite urban residence and hospital access, a subset of patients still sought traditional healers, highlighting that socio-cultural influences rather than geographic access often drive care-seeking decisions.

Perianal pain (84%) and rectal bleeding (50%) were the most frequently reported symptoms in this study, consistent with global literature identifying pain and bleeding as hallmark presentations of hemorrhoids(1). Studies from Ethiopia, such as the University of Gondar outpatient survey, reported rectal bleeding in approximately 40–45% of patients, a lower prevalence than observed in this study, while perianal pain was also common but less dominant(18). In South Asia, hospital-based studies from India documented rectal bleeding in 65–70% of cases and pain in over 70%, reflecting higher prevalence rates in advanced disease presentations(19). Western community-based studies often report lower frequencies, with rectal bleeding in 30–40% and pain in 25–35%, particularly in internal hemorrhoids where pain is less pronounced unless thrombosed(1). A recent global meta-analysis confirmed that bleeding and pain remain the most consistent symptoms across diverse populations, though prevalence varies depending on diagnostic criteria and whether hospital or community cohorts are studied(20). Overall, these comparisons indicate that the high rates of pain and bleeding in this study reflect a hospital-based population with more advanced disease, while still aligning with international evidence that these symptoms are the defining clinical hallmarks of hemorrhoids.

Hospital interventions, particularly hemorrhoidectomy (31.8%), achieved highly favorable results, with 97.3% of patients reporting improvement. This success rate is comparable to international studies where surgical management remains the gold standard for advanced hemorrhoids(1,4). Recent global evidence, including a nationwide cohort from Turkey demonstrating high patient satisfaction and rapid recovery(21), and a randomized trial from India showing reduced pain and shorter hospital stay with closed hemorrhoidectomy(22), further supports the effectiveness of surgical care. The low rate of worsening outcomes (0.2%) underscores the effectiveness of biomedical care.

Despite the availability of hospital care, 6.2% of patients reported seeking traditional healers. The most common modalities included topical application of corrosive chemicals (71.4%), ingestible preparations (39.3%), and pricking with sharp materials (14.3%). Complications were strikingly frequent, with 85.7% of users experiencing adverse outcomes such as persistent perianal pain, anal stenosis, and incontinence. These findings are consistent with reports from North Gondar, where traditional healers similarly relied on herbal preparations and invasive techniques without biomedical validation, often resulting in severe morbidity and long-term disability(18). Comparable studies from Nigeria and Côte d’Ivoire have also documented high complication rates linked to corrosive substances and unstandardized herbal remedies(6,7), underscoring that sociocultural trust in traditional healers continues to outweigh biomedical evidence of harm. Recent Ethiopian qualitative work confirmed similar unsafe practices, including herbal fumigation and corrosive topical agents, with poor outcomes in severe cases(23). Taken together, these results highlight the urgent need for community education and culturally sensitive engagement with traditional practitioners to mitigate unsafe practices and reduce preventable morbidity.

Despite the high complication burden, community attitudes toward traditional treatment remained largely positive or neutral, with nearly half of users (42.9%) reporting favorable perceptions. Decisions to seek traditional healers were strongly shaped by family members (67.9%) and peers (71.4%), underscoring the social embeddedness of care-seeking behavior. This pattern reflects the entrenched role of traditional medicine in Ethiopia, where more than 90% of the population relies on indigenous remedies for primary care(8). WHO’s Global Traditional Medicine Centre(24) similarly reported that 80–90% of populations in developing countries depend on traditional medicine, reinforcing the scale of reliance. Similar findings have been reported in North Gondar, where patients continued to endorse traditional healers despite adverse outcomes(18), and in Nigeria, where sociocultural beliefs and fear of recurrence were major drivers of traditional care use(6). Studies from Côte d’Ivoire likewise highlight how misconceptions and trust in healers perpetuate reliance on traditional practices, even when biomedical services are accessible(7). Taken together, these results suggest that trust in healers, cultural continuity, and limited awareness of biomedical risks contribute to sustained use of traditional remedies, even among urban populations with hospital access. Addressing these perceptions requires culturally sensitive education and engagement strategies that respect community beliefs while reducing unsafe practices.

### Strengths and Limitations

The relatively large sample size and the dual focus on hospital outcomes and traditional treatment complications provide a comprehensive perspective on hemorrhoid care. However, being cross-sectional and hospital-based, the study may underrepresent community cases managed exclusively by traditional healers, and self-reported perceptions are subject to recall bias.

## Conclusion

Traditional hemorrhoid treatment in Southern Ethiopia is associated with high complication rates, yet community perceptions remain favorable due to sociocultural influences. Hospital management demonstrates superior outcomes. Bridging the gap between biomedical care and community beliefs is essential to reduce morbidity and promote safe, effective treatment.

### Recommendations

Community education initiatives should be strengthened to improve awareness of the risks associated with harmful traditional hemorrhoid practices, particularly the use of corrosive chemicals and invasive procedures. Traditional healers should be engaged and trained through culturally sensitive approaches, with integration into public health strategies to minimize unsafe practices. Further community-based research is needed to capture cases managed exclusively by traditional healers and to better understand the sociocultural drivers of care-seeking behavior.

## Data Availability

The dataset generated and analyzed during the current study has been deposited in the Zenodo repository. In accordance with journal policy, the dataset will become publicly accessible upon acceptance of the manuscript

